# Effects of cattle on vector-borne disease risk to humans: A systematic review

**DOI:** 10.1101/2023.02.08.23285683

**Authors:** Sulagna Chakraborty, Siyu Gao, Brian. F Allan, Rebecca Lee Smith

## Abstract

Vector-borne diseases (VBDs) may be transmitted among humans, domestic animals, and wildlife, with cattle in particular serving as an important source of exposure risk to humans. The close associations between humans and cattle can facilitate transmission of numerous VBDs which can impact nations’ public health and economic security. Published studies demonstrate that cattle can influence human exposure risk positively, negatively or have no effect. There is a critical need to synthesize the information in the scientific literature on this subject, in order to illuminate the various ecological mechanisms that can affect the risk of humans contracting VBDs from cattle. Therefore, the aim of this systematic review was to review the scientific literature, provide a synthesis of the possible effects of cattle on VBD risk to humans, and propose future directions for research. This study was performed according to the PRISMA 2020 extension guidelines for systematic review. After screening 470 peer-reviewed articles published between 1999 – 2019 using the databases Web of Science Core Collection, PubMed Central, CABI Global Health, and Google Scholar, and utilizing forward and backward search techniques, we identified 127 papers that met inclusion criteria. Results of the systematic review indicate that cattle can be beneficial or harmful to human health with respect to VBDs depending on vector and pathogen ecology and livestock management practices. Cattle can increase risk of exposure to infections transmitted by tsetse flies and ticks, followed by sandflies and mosquitoes, through a variety of mechanisms. However, cattle can have a protective effect when the vector prefers to feed on cattle instead of humans and when chemical control measures (e.g., acaricides/insecticides), semio-chemicals, and other integrated vector control measures are utilized in the community. We highlight that further research is needed to determine ways in which these mechanisms may be exploited to reduce VBD risk in humans.

**Author Summary:** Vector-borne diseases (VBDs) are caused by infections transmitted by blood-feeding arthropods from an infected to an uninfected organism. These infections may be caused by pathogenic bacteria, viruses, or protozoans and arthropods may transmit these infections to humans, domestic animals, and wildlife. Humans and cattle spend a significant amount of time in close proximity with each other through various activities such as agriculture, animal husbandry, trading, and animal farming, which can potentially increase risk to human health. Previously published studies indicated cattle can impact VBD transmission both positively and negatively, however, there has not been a recent synthesis of the scientific literature on this subject. Through this global systematic review of the scientific literature, we found that cattle could have either harmful or beneficial impacts on human health when it comes to VBDs, but most often increase exposure risk to VBDs in humans. We identified various mechanisms from the scientific literature by which cattle can impact VBD risk in humans. Further research is needed to better understand specific ecological mechanisms by which cattle impact human health and develop measures that will prevent and reduce VBD exposure risk in humans.

## Introduction

Vector-borne diseases (VBDs) are caused by infectious agents that are transmitted by hematophagous arthropods, which can transmit a wide range of pathogens to other organisms (1). Important arthropod vectors of infectious diseases include ticks (*Ixodoidea*), mosquitoes (*Culicidae*), sandflies (*Phlebotominae*), tsetse flies (*Glossinidae*), black flies (*Simuliidae*), and kissing bugs (*Triatominae*). For over a century, vector-borne diseases have been the subject of scientific research because of the severe concern they pose to human and animal health (2 - 5). VBDs account for more than one billion cases, one million deaths, and one-sixth of worldwide disability and illnesses annually (6). Common examples of VBDs include malaria, Lyme disease, Rift valley fever, Chikungunya, West Nile virus and other bacterial, protozoal, and viral diseases (7). Torto and Tchouassi (8) estimate ∼80% of the world’s human population is at risk of exposure to one or more VBDs. Along with their negative impacts on human and animal health, VBDs may have detrimental effects on sustainable development and can cause significant economic losses (9). When a VBD’s prevalence reaches a critical level in a country, human mobility, trade, foreign investment, savings, and land use are all likely to suffer unfavorable consequences (10). As a direct result, VBDs are not only an increasing public health problem, but also have a negative macroeconomic impact on society. Therefore, it is vital to continue efforts to better understand, analyze, and manage health risks due to VBDs and inform effective preventative measures.

Transmission cycles for many VBDs may involve humans, domestic animals, wildlife, and various facets of their environment (11). Environmental factors such as climate may strongly affect the rate of transmission of VBDs; for example, changing temperatures and precipitation due to climate change have been associated with an increase in vector prevalence and transmission (12). VBD exposure risk to humans is especially increased in low-income countries, driven in part by people who are involved in occupations where they handle cattle and other livestock, notably farmers, agricultural laborers, slaughterhouse workers etc. Additionally, people who live in close proximity to cattle or are allied to animal husbandry, as well as those involved in treating and caring for livestock, often are at higher risk of VBDs (13). Socioeconomic disparities have also been associated with increased disease incidence.

Individuals in disadvantaged areas may be unaware of these diseases or associated risk factors, lack access to health facilities and infrastructure, and may follow few to no preventive measures, such as use of insecticide-treated bed nets or vaccines (14). Finally, some tropical VBDs such as leishmaniasis are classified as neglected, and so do not receive sufficient public health attention and funding, complicating disease prevention efforts (15).

Cattle are among the most economically and culturally significant domesticated animals globally (16). There is considerable overlap between cattle and humans through our economic activities, occupations, and cattle being a source of food and recreation (17). Proximity between humans and cattle through agriculture, animal husbandry, and trade provides opportunities for disease transmission. Biotic factors including age, sex, and breed of cattle and their interactions with domestic and wild animals along with abiotic factors such as climate and environmental conditions may influence disease transmission. These factors interact with vector species abundance, longevity, feeding cycle and blood meal host choice as key predictors for how VBD transmission occurs (18).

The overarching role of cattle in the transmission and spread of VBDs in humans is a major gap in our understanding of the ecology and epidemiology of these diseases. Thus, it is important to determine if these human-cattle connections can impact human health via VBDs. Since vectors such as ticks and mosquitoes often feed on multiple host species and may spread various pathogens to humans, livestock, and wildlife, the role of cattle in VBD transmission is complicated. Cattle have various roles to play when it comes to vector-borne disease transmission. For example, they can function as blood meal hosts for arthropod vectors and thereby increase the abundance of vector species (19 - 20), and as reservoir hosts for pathogens and thereby increase pathogen prevalence (21 - 22). In contrast, cattle can be more attractive to biting vectors than humans and thereby act as shields against vector bites to humans preventing pathogen transfer in specific circumstances (23 - 24).

Through this systematic review, we have been able to identify multiple VBDs for which cattle have a direct or indirect role in infection transmission; however, for many VBDs the exact role of cattle in the ecology of these diseases remains undetermined. Arguably the most contentious matter has been that of the zoo-prophylactic role of cattle (i.e., cattle acting as a barrier against disease transmission by absorbing vector bites and thereby having a protective effect for humans). There is a significant debate in the scientific literature over whether the presence of cattle near humans can substantially reduce disease incidence. There are studies that have both supported and failed to support the zoo-prophylactic effect of cattle on VBD risk to humans, such as in the case of malaria (25-26).

Globally, there are numerous studies that directly investigate whether cattle increase or decrease risk of VBD exposure in humans, and yet currently there is no synthesis of the existing information on this subject. This gap can impact public health and epidemiological measures that countries can take to prevent VBD transmission. Thus, our goal was to conduct a systematic review of the scientific literature to synthesize and present the findings on the conditions by which cattle increase or decrease human risk of exposure to VBDs and by what ecological mechanisms.

## Methods

### Search strategy

We conducted a systematic review of the published scientific literature to determine whether studies report a positive, negative, or neutral impact of cattle on human exposure risk to vector-borne diseases (i.e., whether cattle increased, decreased, or had no effect, respectively). Following the PRISMA 2020 (Preferred Reporting Items for Systematic Reviews and Meta-Analyses) guidelines (27), we developed a search algorithm that would enable us to extract scientific papers on this subject from various databases, using the following search string:

TS = (cattle AND (tick-borne illness OR tick borne disease OR mosquito-borne illness OR mosquito borne disease OR vector-borne illness OR vector borne disease) AND human health) AND LANGUAGE: (English), year range 1999 to 2019.

The initial step after developing the search string was to check if there were other systematic reviews on this topic through the Cochrane Database of Systematic Reviews. We used the keyword search comprising “cattle AND vector-borne diseases AND human health” on Cochrane, which did not yield any systematic review; it did yield one article (28) which was outside the scope of this study. Subsequently, we used the following databases to execute the search string to identify relevant articles: CABI Global Health, Web of Science Core Collection, PubMed Central and Google Scholar. Articles were also identified through forward and backward searches from citations in both included and excluded articles. Titles and abstracts of the articles identified through keyword search, forward, and backward searches were screened against the study inclusion criteria. Potentially relevant articles were retrieved for evaluation of the full text and duplicates were removed.

Full text articles were further screened and evaluated using the full study inclusion criteria, which were: a) study should be published between 1999 - 2019 to ensure recency; b) study language should be in English; c) study incorporates all of the key terms: vector (specifically searched for the terms tick-borne, mosquito-borne), vector-borne disease, humans, and cattle; d) study explores a possible connection between vector, cattle, and human; e) full texts of the articles available (full texts were accessed through University of Illinois library, Google Scholar and World Cat database). Articles were excluded from the study if they met any of the following exclusion criteria: a) analysis excludes vector-borne diseases; b) study fails to mention a vector arthropod that transmits disease; c) study does not include cattle involvement; d) control studies, vaccine studies, therapeutic studies, or review papers; e) studies involving only experimental lab infection; f) studies whose full texts could not be accessed after multiple attempts from various sources; g) non-English language articles; h) studies that only include pathogens that do not cause human infection; i) studies conducted outside our time period. This review had no geographical restrictions.

Two reviewers independently assessed inclusion and exclusion criteria. A third reviewer assessed studies for which the reviewers disagreed. We calculated Cohen’s Kappa statistic to estimate inter-rater agreement between the first two raters. Article search was conducted between September 30, 2019, until June 8, 2020.

### Data extraction and synthesis

Methodological and outcome variables from each selected study were collected in a database, including article title, authors, publication year, country, study type, vector taxa, main implications of each study, and database source for this review. We summarized the common themes and findings of the included studies narratively. To better illustrate the impact of cattle on human health due to VBDs, we characterized the effect of cattle on VBD exposure risk to humans into three categories: a) neither beneficial nor harmful (no association), b) beneficial, or c) harmful, and identified any mechanisms investigated or proposed by the authors that contributed to human risk of exposure to VBDs by cattle.

### Study quality assessment

To evaluate the quality of the included studies, we used The Strengthening the Reporting of Observational Studies in Epidemiology (STROBE) Statement guidelines for reporting observational studies (29). The first two authors independently rated each included study. These articles were rated on a score ranging from 0 - 2, depending on whether the criteria were unmentioned or < ¼ met (0), ¼ - ¾ met (1), or > ¾ met (2). The following were the critical criteria used in rating the study quality: (a) was the research question clearly stated? (b) what was the study design and study setting used? (c) what was the sample size used? (d) were the subjects in the study representative of the target population? (e) were the main findings clearly described? (f) were there any confounding variables? (g) did the researchers use appropriate analytical methods? and (h) was the study located in the predefined area of interest? The study quality score measured the strength of study evidence for reporting here, but studies were not excluded based on quality.

## Results

### Study selection, Inter-rater agreement & Study quality assessment

We screened 470 articles through a keyword search using our search algorithm. The number of articles identified from each database, and number of articles included and excluded in our review, are listed in Fig 1. After removal of 12 duplicates, we identified 458 unique articles. Out of these, 331 did not meet the inclusion criteria. The remaining 127 articles are the final pool of studies included in this review. S1 Appendix provides the complete reference list of all studies that were finally included after performing the database search. Cohen’s Kappa statistic for inter-rater agreement between two raters was calculated to be 0.835, indicating strong agreement between the article reviewers (30). Averaging the ratings provided by two members who conducted the study quality assessment, the reviewed studies averaged a score of 1.22 out of 2, with a standard deviation of 0.55 for 127 articles.

**Fig 1:**
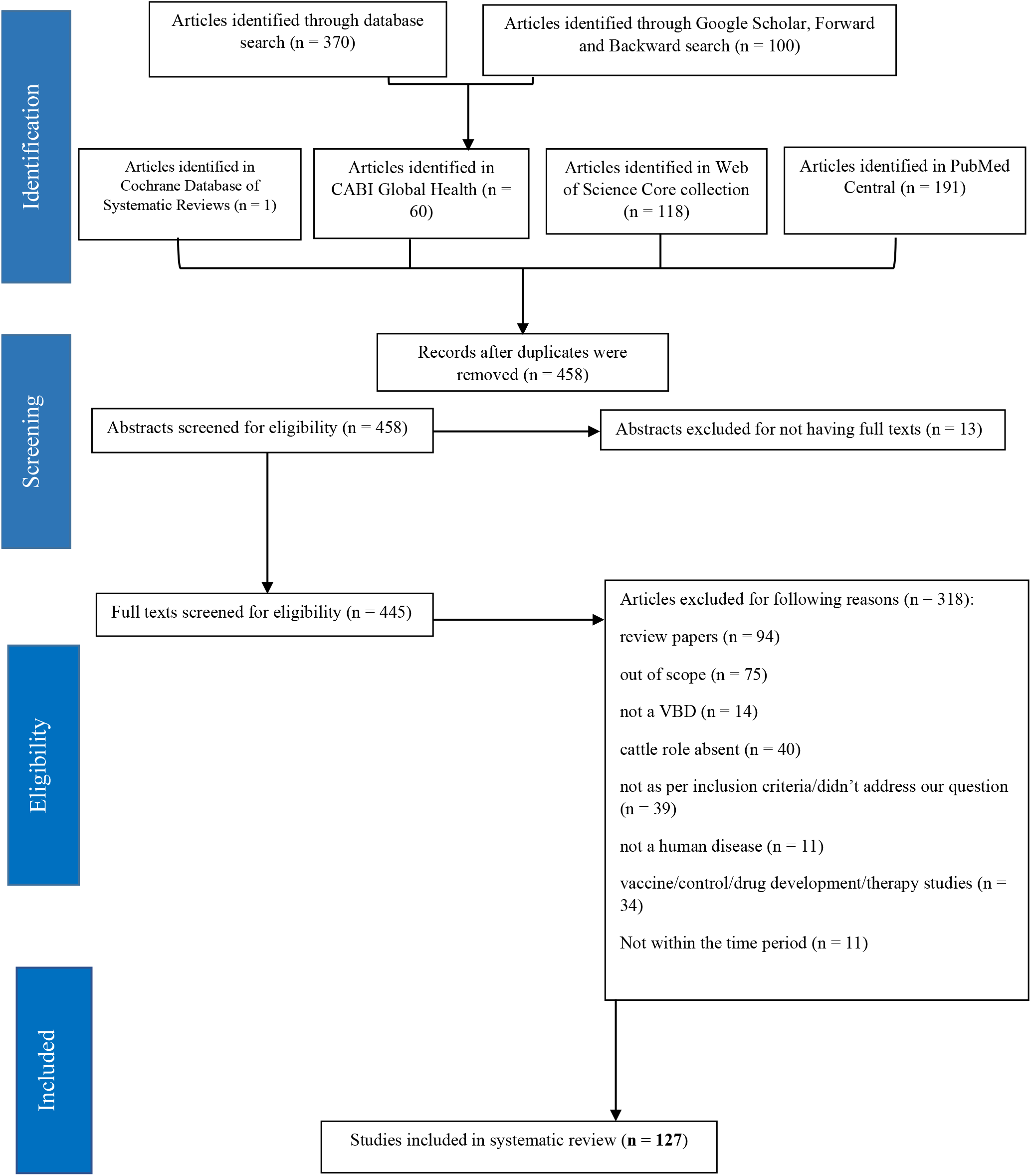
Flowchart for the systematic review process.

### Study characteristics

The most common study methods used by researchers worldwide were observational studies, studies that utilized molecular biology, phylogenetic, and genetic techniques, mathematical modeling studies, and entomological studies. Several studies involved more than one research method or study design to collect various kinds of data from the study population. Articles were included from 69 individual countries and the European Union. Of note, only one study from the United States was in the final pool of included studies. The majority of the papers included in this review were from the African continent (N = 82), followed by Asia (N = 37), Europe (N = 24) and only a few papers from South America (N = 4), North America (N = 4) and one study from Oceania/Australia (N = 1; Fig 2). Note that there were several studies that were based in more than one country; for example, if a study was conducted both in Cameroon and Nigeria, it was counted twice in Fig 2.

**Fig 2:**
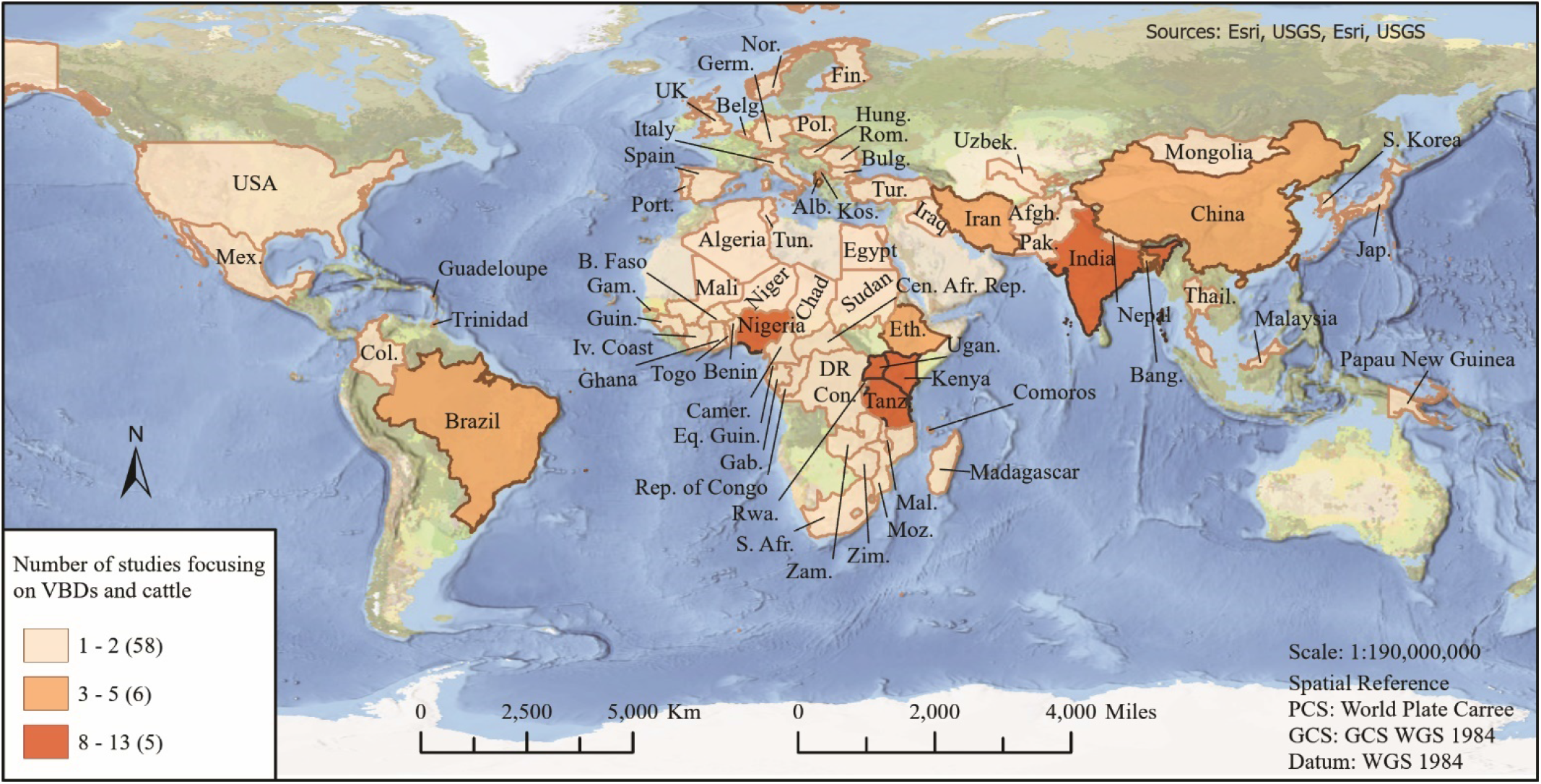
World map representing countries and number of studies included in this review.

Based upon the findings reported by study authors, effects of cattle on human health with respect to exposure to VBDs were divided into three categories: beneficial, harmful, or neither beneficial nor harmful (no association) (Fig 3, Table 1). The most beneficial impact of cattle was observed in the case of infections spread by mosquitoes and sandflies. Cattle sometimes had beneficial impact when it came to tick-borne diseases, especially in integrated cattle and wildlife communities. In such communities, when cattle were treated with acaricides they reduced the abundance of host-seeking ticks in the environment, thereby reducing tick-borne disease risk for wildlife and humans (31-32). However, effects of cattle on tick-borne disease risk were sometimes harmful as well. Cattle have been found to be a major risk factor for humans when it comes to diseases spread by tsetse flies. There were some studies for each vector taxon that stated that cattle had neither harmful nor beneficial exposure impacts on human health, with the exception of tsetse flies. From this systematic review, we found that in the case of six VBDs and in the case of few tickborne pathogens, cattle impacted VBD exposure risk in humans both positively and negatively, for 14 major VBDs cattle were harmful and for 2 VBDs cattle were beneficial (Table 1). We also identified various mechanisms from these published studies by which cattle can positively or negatively impact human exposure risk to VBDs (Table 2). Seven mechanisms were identified by which cattle may impact VBD exposure risk in humans. Overall, we find that cattle tend to increase the risk of exposure to VBDs in humans, but there are circumstances when cattle can reduce or have no effect on VBD exposure risk as well.

**Table 1:** Role of cattle on risk of exposure to vector-borne diseases in humans from studies included in this review.

| Vector-borne disease | Vector | Role of cattle | Selected Sources |
| --- | --- | --- | --- |
| Rift valley fever<br>(RVF) | Mosquitoes (primary vector-*Aedes* species, secondary vector *Culex* species) | Harmful. (See Table 2: 4 - 6) | 33 - 35 |
| Malaria | Mosquitoes (female *Anopheles* species) | Both harmful and beneficial effects observed. (See Table 2: 1, 4, 7) | 18, 24, 36, 37, 38, 39, 40, 41 |
| Japanese Encephalitis<br>(JE) | Mosquitoes (*Culex* species) | Beneficial and potentially harmful effects observed. (See Table 2: 1, 3) | 42-43 |
| Chagas disease | Kissing bug (*Triatoma brasiliensis brasiliensis*) | Potentially harmful (See Table 2: 1, 4) | 44 -45 (Not part of included studies) |
| West Nile Virus<br>(WNV) | Mosquitoes (*Culex* species) | Possibly harmful but in rare instances (debatable) (See Table 2: 4) | 46-47 (Not part of included studies) |
| St. Louis Encephalitis (SLE) | Mosquitoes (*Culex* species) | Potentially harmful (See Table 2: 4) | 48 |
| Human African Trypanosomiasis (HAT) or sleeping sickness | Tsetse fly (*Glossina* species) | Harmful & Beneficial. (See Table 2: 4-7) | 49, 50, 51, 52 |
| Lyme Disease | Ticks (*Ixodes scapularis*, *Ixodes pacificus*, *Ixodes ricinus*) | Beneficial (See Table 2: 2 - 3) | 53-54 |
| Crimean-Congo hemorrhagic fever (CCHF) | Ticks (primary vector *Hyalomma* species; secondary vector *Rhipicephalus*, *Haemaphysalis* and *Dermacentor* species) | Harmful and Beneficial (See Table 2: 4 - 5, 7) | 31, 32, 55, 56, 57, 58, 59. |
| Anaplasmosis | Ticks (*Ixodes scapularis*, *Ixodes pacificus*, *Ixodes ricinus*) | Harmful (See Table 2: 4, 6) | 60, 61, 62, 63 |
| Babesiosis | Ticks (*Babesia divergens*; *Babesia microti*, *Babesia bovis*, *Babesia bigemina*) | Harmful (See Table 2: 6) | 64, 65, 66 (Not all papers are part of included studies) |
| Dugbe virus | Ticks (*Hyalomma*, *Amblyomma* and *Rhipicephalus* species) | Harmful and Beneficial (See Table 2: 4, 7) | 67; 68, 69<br>31, 32 |
| Thogoto virus | Ticks (*Rhipicephalus praetextatus*) | Potentially beneficial (See Table 2: 7). | 31, 32 |
| Alkhurma/Alkhumra hemorrhagic fever (AKHV) | Ticks (*Ornithodoros savignyi*, *Hyalomma dromedari*) | Harmful (See Table 2: 5) | 70, 71, 72 (Not all papers are part of included studies) |
| Kyasanur Forest Disease (KFD) | Ticks (*Haemaphysalis spinigera*, *Haemaphysalis turturis*) | Harmful (See Table 2: 4 - 6) | 73, 74, 75, 76 (Not all papers are part of included studies) |
| Ehrlichiosis | Ticks (*Amblyomma americanum*, *Ixodes scapularis*) | Harmful (See Table 2: 4) | 62, 77 |
| Tickborne Encephalitis Virus (TBEV) | Ticks (*Ixodes ricinus*, *Ixodes persiculatus*, *Haemaphysalis punctata*, *Dermacentor marginatus*) | Harmful (See Table 2: 4, 6) | 78, 79 |
| African tick bite fever (ATBF) | Ticks (*Amblyomma variegatum*, *Amblyomma hebraeum*) | Harmful (See Table 2: 4) | 80, 81, 82 (Not all papers are part of included studies) |
| Rocky Mountain Spotted Fever (RMSF) | Ticks (*Dermacenter variabilis*, *Amblyomma americanum*, *Rhipicephalus sanguineus*, *Amblyomma cajennense*) | Harmful (See Table 2: 4) | 83 |
| Spotted fever group<br>(SFG) rickettsioses | Ticks (*Amblyomma maculatum*, *Rhipicephalus*, *Dermacentor*, *Hyalomma* and *Ixodes* species) | Harmful (See Table 2: 4) | 22, 80, 84 |
| Q fever | Ticks (*Dermacentor* species, *Hyalomma* species, *Haemaphysalis* species, *Rhipicephalus* species, *Ixodes* species) | Harmful (See Table 2: 4 - 5) | 85, 86 |
| Cutaneous Leishmaniasis and Visceral Leishmaniasis (Kala azar) | Sandfly (*Lutzomyia gomezi*, *Lutzomyia longipalpis*, *Lutzomyia ovallesi*, *Phlebotomus argentipes*, *Phlebotomus papatasi* *Phlebotomus sergenti*, *Sergentomyia squamipleuris*) | Both harmful and beneficial effects observed. (See Table 2: 2, 4, 7) | 87, 88, 89, 90, 91, 92, 93 |

**Table 2:**
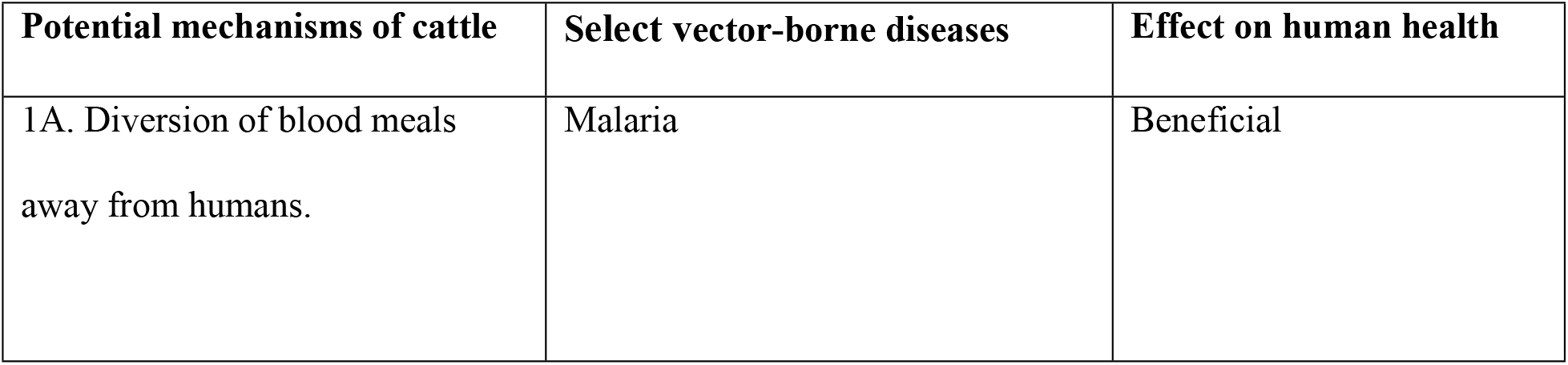
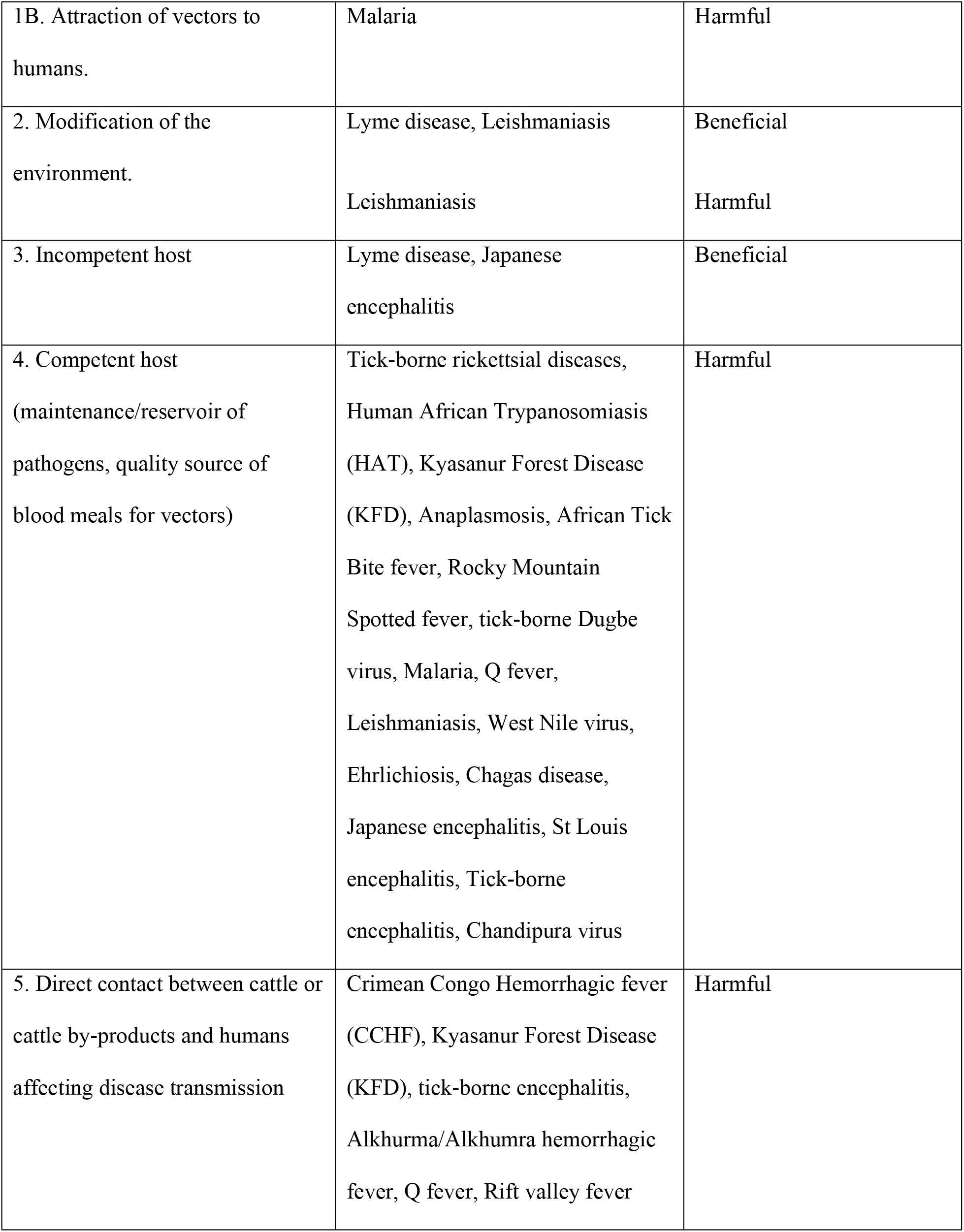
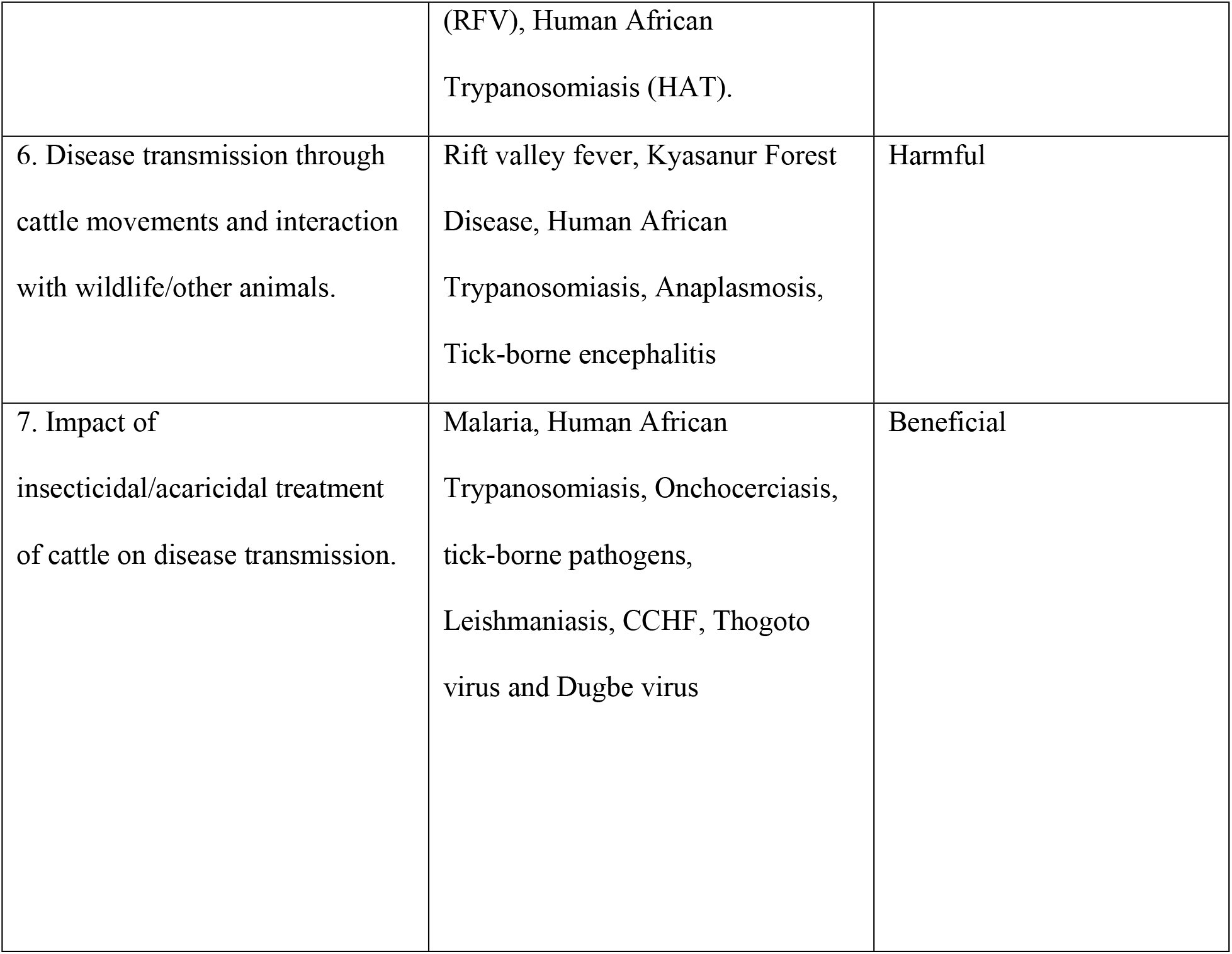
Mechanisms identified from included studies by which cattle impact vector-borne disease exposure risk in humans.

**Fig 3:**
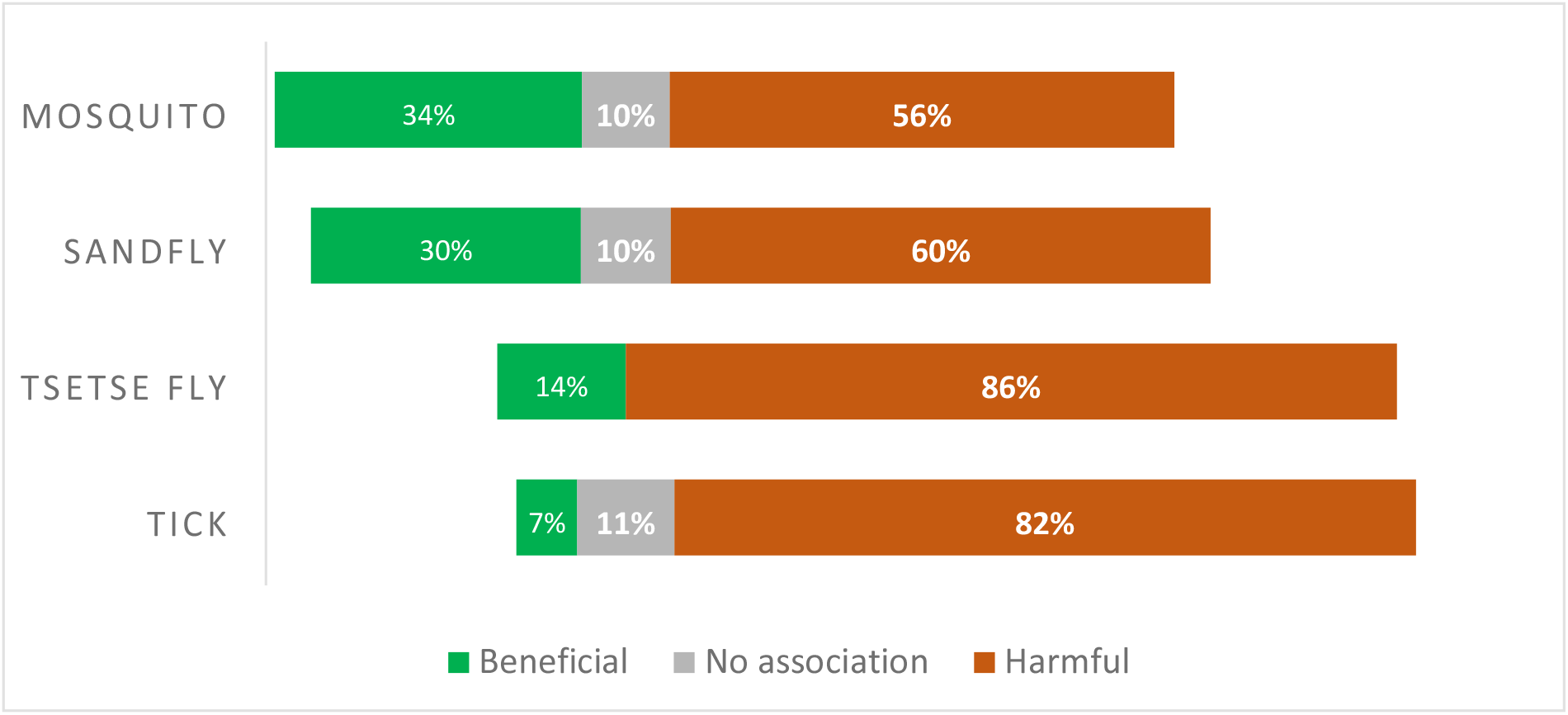
Cattle impact on risk of exposure to vector-borne diseases on human health, divided into beneficial, harmful and no effect by major vector taxon, covered in this review.

## Discussion

Cattle and other livestock animals are principal elements in agriculture, animal husbandry, trade, economic activities as well as in cultural practices of people around the world. People who are in close contact with cattle due to employment, commerce or for traditional reasons often are at higher risk for contracting various vector-borne and zoonotic diseases (13, 94).

We identified a critical gap in the scientific literature about the various roles cattle can play in vector-borne disease transmission. We systematically reviewed articles from the scientific literature to synthesize the available information to better understand how cattle impacts VBD exposure risk in humans. We categorized the impacts of cattle on VBD exposure risk in humans by effect (positive, negative, neutral) and by vector taxa (mosquitoes, sandflies, ticks, tsetse flies). We also identified seven mechanisms by which cattle positively or negatively impact VBD exposure risk in human health.

Research on this topic has been concentrated geographically in countries located in sub-Saharan Africa and southern Asia. Cattle appear to have both beneficial and harmful impacts on human health with respect to VBDs, but they tend to overwhelmingly increase risk of exposure to VBDs in humans. These effects of cattle on VBD exposure risk in humans depends on various ecological conditions, on the vector taxa along with other environmental factors.

In order to understand the mechanisms by which cattle can impact risk of human exposure to vector-borne diseases we define some key terms here, which are sometimes used inconsistently in the literature we reviewed. A reservoir is defined as one or more epidemiologically connected populations of host species in which the pathogen can be permanently maintained and from which infection is transmitted (95). A maintenance population can be defined as a host population in which a pathogen persists because the population size is greater than the critical community threshold (95, 96). An amplifying host is an organism in which an infectious agent (such as a virus or bacterium) that is pathogenic for some other species is able to replicate rapidly and to high concentrations (97) as evidenced in the case of Japanese encephalitis virus for which pigs are the amplifying host species (98). The ability to obtain and transmit pathogens to other organisms refers to the competence of the host in transmitting that infection (99).

In our review and synthesis of the studies included in this systematic review, we identified seven classes of mechanisms by which cattle can impact risk of exposure to vector-borne diseases in humans, which are discussed below. We also recorded the number of times the papers within our included studies invoked one or more of these mechanisms.

### Mechanism I: Diversion and attraction of vector blood meals

There is considerable evidence in the published literature to support scenarios in which there is a zoo-prophylactic effect of cattle on human health. Zoo-prophylaxis occurs when the presence of cattle can function as a barrier against potential vectors and as an alternative host that deflect blood meals away from humans. This phenomenon has been observed and studied greatly in the context of malaria (24, 26, 39, 100, 101). This mechanism was observed 16 times from the included studies. Contrastingly, there is also evidence of instances when zoo-prophylaxis has not been observed (25, 39, 101, 102, 103, 104, 105, 106, 107). Multiples studies indicate that certain requirements/conditions need to be present for zoo-prophylaxis to occur (24, 26, 38). In the case of malaria specifically these conditions are a) zoophilic and exophilic vector, b) habitat separation between human and host animal quarters, and c) augmentation of zoo-prophylaxis with insecticide treatment of animals or co-intervention of long-lasting insecticide-treated nets and/or indoor residual spraying. Presence or absence of these requirements might explain to a certain extent why cattle may or may not always be observed to be zoo-prophylactic.

The opposite of zoo-prophylaxis is zoo-potentiation, whereby livestock contribute to an increase in VBD transmission by attracting vector bites to humans; this often occurs where livestock are housed within or near human sleeping quarters and for vector species that prefer human hosts, such as with some species of mosquitoes that transmit human malaria (24). As opposed to zoo-prophylaxis, the improved availability of blood meals by increasing the presence of cattle increases mosquito survival, which counters the beneficial impact of diverting blood meals on endemic and epidemic malaria (104). Tirados et al (101) showed in field studies that in outdoor conditions, cattle had no prophylactic effect on humans but the presence of cattle outside with humans indoors had some protective effect. Clearly the mere presence of cattle may not always be sufficient to protect humans from malaria-carrying mosquitoes. Cattle may attract more vectors (108), and they can increase local abundance of specific vectors causing both cattle and human VBDs such as tick-borne pathogens, malaria and leishmaniasis. We observed this mechanism within our included studies 18 times.

### Mechanism II: Modification of the environment

A second mechanism by which cattle may impact VBD risk is through physical modification of the environment. Cattle may be able to modify the environment making it either suitable or unsuitable for certain vectors thereby impacting VBD exposure. For example, cattle can modulate the risk for Lyme disease by reducing the prevalence of questing vector ticks in a managed pasture (53). In this study, cattle modified the vegetation by their grazing thereby rendering the microclimate more arid and making the environment less suitable for the survival of ticks. Another instance where cattle may have a beneficial impact on human exposure is the case of Leishmaniasis: Bern et al (88) found that household cattle ownership was associated with lower risk of contracting the infection and presence of large numbers of cattle around houses had a protective effect. Conversely research by Singh et al (92) found that one of the primary vectors of Leishmaniasis in India, *Phlebotomous argentipes*, preferred to mate in cattle sheds and in soils that were more alkaline than in human houses, whereas another vector *P. papatasi* preferred the soil of human houses with neutral pH. This is another example where cattle may modify environments to be more suitable for vector survival and where it can have a negative and positive effect on human health depending on ecological attributes of the vector. We found only 6 instances when this mechanism was discussed in our pool of studies.

### Mechanism III: Incompetent host

Cattle have been found to be an incompetent reservoir host species for certain pathogens such as the causative agent for Lyme disease (54). Non-infected ticks that feed on cattle fail to acquire spirochetes, but also infected ticks may even lose their infection during the course of blood meals from cattle (53). Similarly, Samuel et al (43) reported that a decrease in the cattle-to-pig ratio might be one of the reasons for an increase in Japanese encephalitis virus (JEV) infection among children in India. Pigs are competent reservoir hosts for JEV, whereas cattle are a dead-end host for JEV, and presence of cattle may have a protective effect on humans. This mechanism was only observed 3 times within our pool of studies.

### Mechanism IV: Competent host

In other cases, cattle may serve as competent reservoirs, maintenance hosts, quality source of bloodmeals or amplifying hosts for several vector-borne diseases. In some circumstances, cattle can maintain vector-borne pathogens in their system that remain undetected, which may cause an outbreak when the right conditions arise (109). For example, in the case of Human African Trypanosomiasis (HAT), cattle along with pigs serve as reservoirs of human infective *Trypanosoma brucei rhodesiense* and also serve as blood meal hosts for the tsetse fly vector (49, 110). Cattle have been found to harbor all life stages of ticks that can transmit Kyasanur Forest Disease (KFD) to humans and have also found to maintain a low level of the KFD virus infection without succumbing to the disease (73, 75, 111). Cattle are reservoirs for the pathogens that cause Anaplasmosis and can be co-infected with two or more *Anaplasma* species simultaneously (61, 112, 113). Cattle can be reservoirs for several species of tick-borne rickettsial pathogens as well (22, 84). Cattle are also known to maintain tick-borne Dugbe virus in the environment, which primarily affects children (68, 69). Cattle and other ungulates are important reservoirs of the causative agent of Q fever, *Coxiella burnetii* (114, 115). This was the most commonly observed mechanism (found 63 times) in our systematic review and appears to be a better studied mechanism than some of the other mechanisms mentioned here.

### Mechanism V: Direct contact between cattle or cattle by-products and humans affecting disease transmission

Several sources indicate that consumption of dairy products from infected cattle (i.e., after they have been bitten by ticks) or consumption of infected meat itself are risk factors for diseases transmitted by vector arthropods, such as Crimean Congo hemorrhagic fever (CCHF), tick-borne encephalitis, Rift Valley Fever (RVF), and Alkhurma/Alkhumra hemorrhagic fever (57, 70, 78, 116, 117). Handling both live and dead infected cattle and contact with raw animal skins and body fluids of infected cattle can also be risk factors for CCHF and RVF (59, 118, 119). Various studies have also shown that people working in professions in close contact with cattle, such as veterinary professionals, abattoir workers, butchers, farm workers, livestock handlers, traditional pastoralists, tannery workers, and human health professionals, are at risk of contracting CCHF, RVF, HAT, Q fever and other VBDs, either through direct contact with cattle or indirectly via vector bites on the cattle (33, 55, 57, 28, 116, 118, 120, 121). In addition, congregations of large herds of cattle with humans due to trade and religious festivals at trading posts, live animal markets, quarantine facilities, and in slaughterhouses, allow for more opportunities for VBD transmission such as in the cases of RVF and CCHF (56, 122, 123, 124). This is the second most commonly observed mechanism in the reviewed literature, with a count of 28 times.

### Mechanism VI: Disease transmission through cattle movements and interaction with wildlife/other animals

Movement of animals from disease-endemic to non-endemic places and the interaction between cattle and other animal species during grazing activities can also result in geographic spread of VBDs to new foci (117, 124, 125). Omondi et al (62) state that wildlife translocations from areas with vector presence to areas without vector presence can also lead to VBD transmission. Diseases that typically are rare in humans, such as babesiosis, have been found to increase due to dissemination of pathogens through cattle movement (66). Movement of otherwise free-ranging cattle to forest and back into villages have been hypothesized to be a risk factor in the spread of Kyasanur Forest Disease in India (75, 76, 126). Rutto et al (49) showed that in areas where untreated cattle, humans and other livestock come into contact with each other, especially during dry periods at watering points, there can be risk of bovine and human trypanosomiasis transmission. Murase et al (127) demonstrated that cattle might be contracting *Anaplasma phagocytophilum* from contact with wildlife and could easily transmit it to humans in Japan. Similar to mechanism V, anthropogenic activities, and areas such as trading posts, wet markets, religious festivals etc. where there are congregations of cattle, humans, and other domestic or wild animals; these afford opportunities for spillover of pathogens from one species to another (122, 123, 128, 129). We observed this mechanism 26 times in our included pool of studies.

### Mechanism VII: Impact of insecticidal/acaricidal treatment of cattle on disease transmission

A major beneficial impact of cattle on VBD exposure risk in humans is through treatment of cattle with insecticides/acaricides. There is research that shows treatment of cattle with insecticides is associated with a significant decrease of malarial vectors in the environment, when used in conjunction with insecticide-treated bed-nets, indoor residual spraying, and other vector control approaches (37, 38, 104, 130, 131, 132, 133, 134). When cattle are treated with insecticides against tsetse flies or trypanocides against the parasites, it reduces the abundance of vectors and parasites thus preventing transmission of HAT (49, 50, 51, 135). Insecticidal or acaricidal treatment of cattle also has positive impacts in controlling other mosquito vectors (136), blackflies (137), leishmaniasis (138, 139, 140) and preventing tick-borne diseases in non-integrated ecosystems (141, 142).

Acaricidal treatment of cattle can also have beneficial impacts in wildlife and livestock integrated communities. Allan et al (31) and Keesing et al (32) describe that in such integrated communities in Kenya, treatment of cattle with acaricides can reduce abundance of host-seeking ticks in the environment. This could also improve the health of wildlife, domestic animals and humans that co-occur in such shared ecosystems. Treatment of the cattle with specific acaricides reduced the abundance of host-seeking nymph and adult life stages of several tick species (vectors of diseases such as CCHF, Thogoto virus and Dugbe virus), thereby reducing potential for disease transmission. Interactions between cattle and wildlife can have important epidemiological consequences. For example, Ruiz-Fons et al (143) found that in game reserves in Spain where cattle and ungulates coexist, cattle abundance influenced the prevalence of *B. burgdorferi* sensu lato and *A. phagocytophilum* in *I. ricinus* nymphal ticks. Increasing abundance of cattle seemed to increase the risk of other hosts becoming infected by *A. phagocytophilum*, while reducing the risk of becoming infected by *B. burgdorferi* sensu lato. We observed this mechanism in our included pool of studies 17 times.

Treatment of cattle with insecticides either topically or through ingestion can reduce the circulating parasites in the environment along with the targeted vector species, thereby reducing disease burden and vectorial capacity of the vectors. However, efficacy of insecticide treatment of cattle is dependent not just on the feeding preferences of the vector but also on behavioral adaptations of vectors, potential development of resistance among the vectors and parasites, and potential negative consequences for the environment (41, 132, 144, 145, 146). If the targeted vector species in an area are all anthropophilic, then there has to be a multi-pronged approach to control the vectors and VBDs in that area. We summarize this intricate interaction between vector feeding preference and insecticide treatment of cattle on human health in Fig 4.

**Fig 4:**
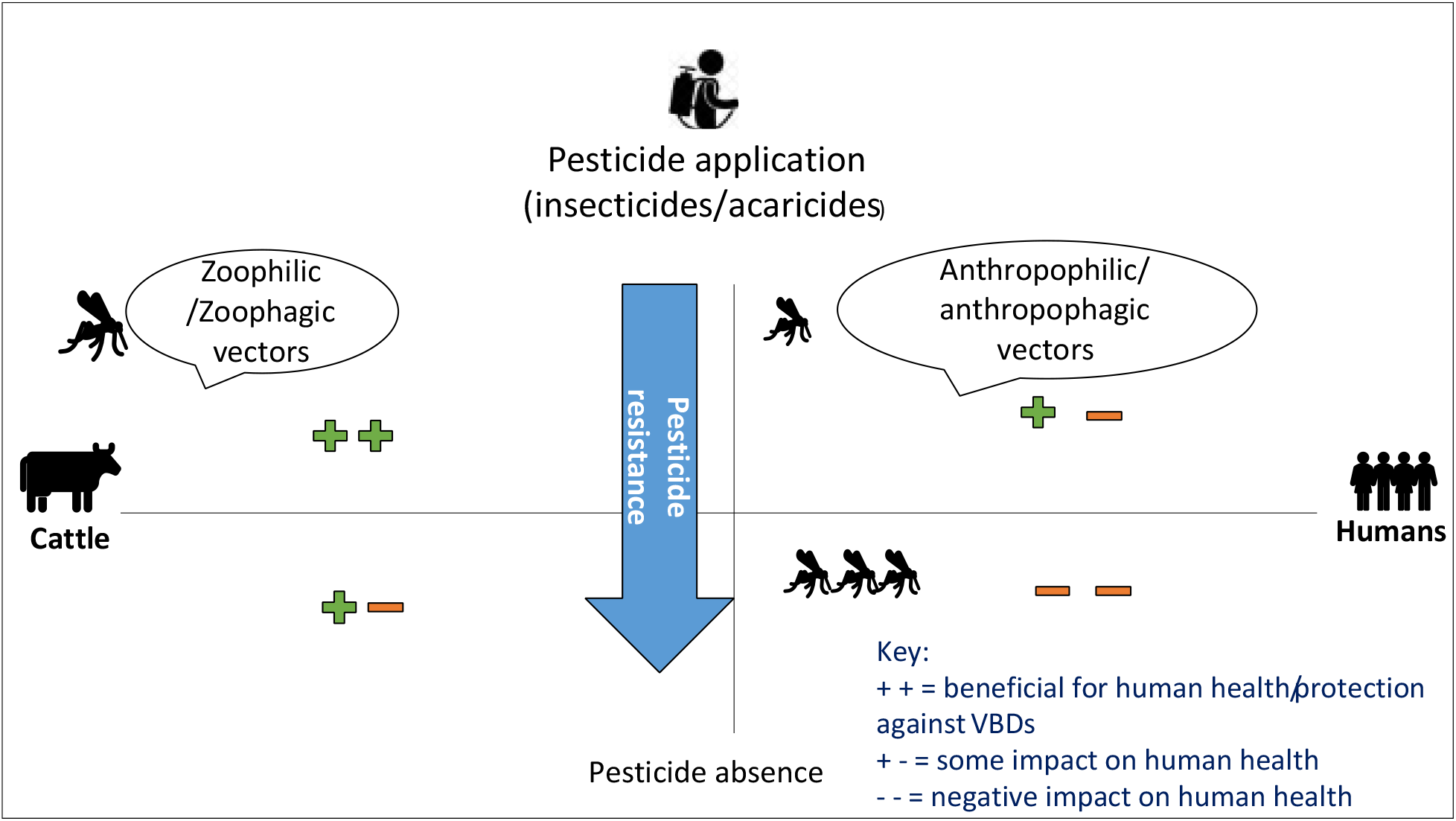
Proposed effects of insecticide-treated cattle and vector feeding preference on human exposure risk.

We hypothesize that when cattle are treated with insecticides and/or vectors preferentially feed on cattle, cattle likely reduce VBD exposure risk in humans by deflecting vector blood meals away from humans and/or reducing the abundance of vectors in the environment. Conversely, when vectors prefer to feed on humans and/or cattle are not treated with insecticides, cattle are likely to increase VBD exposure risk in humans by contributing to an increase in vector abundance, attracting vectors to feed on humans, and/or serving as pathogen reservoir hosts that can transmit the infection to vector arthropods.

Treatment of cattle with various insecticides to prevent disease transmission has been shown to be a critical step in vector control. However, factors such as inadequate market infrastructure, poor awareness, expensive nature of treatments, and local policy enforcement failures can lead to low rates of cattle treatment (49, 125). There is research underway to investigate alternative methods that can be used to control vectors and thereby vector-borne diseases, such as the use of plant-based odor baits (147) and semio-chemicals.

Semio-chemicals are organic compounds that function as signals and enable intra- and inter-specific chemical communication (148). The information conveyed is used for modulating physiological and behavioral activities through the olfactory and taste system (148). Mosquitoes use a variety of sensory cues to find their prey which can differ depending on the specific life stages of the mosquito (149). Various semio-chemicals have been identified that mosquitoes use during oviposition, mating, sugar feeding and host-seeking (149). When semio-chemicals are applied to cattle and livestock, they can attract specific vectors to the cattle and then kill the vectors (150); or they can disrupt mating in the vector (151); additionally, they can be used to repel vectors from finding their preferred hosts (e.g., humans) (151). The search is on for other alternative methods or compounds that can be used in vector control methods that might be less environmentally harmful. For instance, Singh et al (92) evaluated application of plant products at potential sandfly breeding sites to reduce soil pH that might help in preventing vector mating and can be a useful alternative to chemical insecticides for sandfly control/management. Other vector control approaches being developed include gene drive technology (152), infection of *Aedes* mosquitoes with Wolbachia to prevent VBD spread (153, 154), and a variety of environmental modifications (154).

### No effect/no association of cattle on VBD risk

There were some papers that met our inclusion criteria and yet did not explicitly study the impact of cattle on human exposure risk to VBDs (155, 156). In addition, other studies found unclear associations between cattle and VBD risk to humans (157, 158; 159, 160, 161, 162). As we found few such papers in our pool, this indicates the need for diversity of study designs and research methodologies that might better investigate the impact of cattle on VBD exposure risk in humans.

### Limitations

It is important to consider study limitations. Since we opted to focus on more recent papers, the final pool of studies did not include papers published before 1999, which may overlook older but still important research. There may be articles published on this topic in languages other than English which we could not include, and there have been review articles published that may have included otherwise unpublished data which we did not include. In some articles it was difficult to parse out effects of cattle from effects of other livestock since some studies group cattle as part of multiple livestock species, despite our specific use of the term ‘cattle’ instead of ‘livestock’ in our search algorithm. Of note, since this systematic review encompasses information from multiple fields such as ecology, epidemiology, parasitology etc., certain terms/jargons and concepts are interchangeably used making this a challenging question to answer. Finally, our evaluation of mechanisms by which cattle affect VBD exposure risk in humans was dependent on mechanisms invoked by the authors of these studies, some of which were supported by experimental evidence, but others based on field observations or informed opinions.

### Future recommendations and research

The role of cattle in vector-borne disease transmission can be complicated and can prove to be beneficial or harmful in the context of specific VBDs and in specific settings. We recommend future studies explicitly study the various mechanisms by which cattle impact vector-borne disease transmission, as more than one mechanism may operate in specific environmental contexts. We encourage researchers to use a wider variety of study designs than just modeling, serology, molecular analyses, cross-sectional, and retrospective methods. Apart from human and animal health, other factors such as cultural practices, societal norms, age, sex, occupation, human activities and behaviors, and seasonality can predispose individuals to vector-borne diseases. More research is needed to investigate all these factors as well as identify situations in which cattle can be zoo-prophylactic beyond the well-studied example involving human malaria. Since published studies tended to be concentrated from a few specific regions, more research and funding on this topic from other geographic areas (e.g., North America) might yield interesting results. The results from this research could inform public health measures globally to prevent and reduce vector-borne disease transmission. Policy measures, increased funding, and public awareness are all critical steps in the fight against vector-borne diseases since many vectors are opportunistic and can parasitize many different host species.

## Conclusion

The goal of this systematic review was to determine the impact of cattle on human health with respect to vector-borne diseases. Our results show that cattle often increase VBD exposure risk in humans but there is evidence to show that cattle can have a beneficial impact on human health as well. We hypothesized seven mechanisms from the literature through which cattle can impact VBD exposure risk in humans; these mechanisms are dependent on ecological conditions, vector taxa and other environmental factors. In addition, some mechanisms are less studied than others and require further investigation. Hence, it is critical for future studies to delve deeper into the many ways cattle, humans, wildlife, and vectors interact in the environment and develop holistic measures that can be used to protect humans and animals from VBDs as well as prevent negative side effects on the environment.

## Data Availability

S1 Appendix in the supporting information section lists the final pool of included studies that we obtained after performing the article search on databases and screening them against our inclusion and exclusion criteria. This appendix has the names of the papers, author, publication year, country of publication, vector taxa, study implication and database source.

## Acknowledgements

We would like to thank Kelli Trei, Biosciences Librarian and Associate Professor at the University of Illinois Urbana-Champaign, who helped us in the study search process.

## Supporting information

**S1 Appendix. This is the list of the final pool of included studies in this systematic review**

**S1 Table. Completed PRISMA 2020 checklist**

